# Scalable and Resilient SARS-CoV-2 testing in an Academic Centre

**DOI:** 10.1101/2020.04.19.20071373

**Authors:** On behalf of the CRICK COVID-19 Consortium, J Aitken, K Ambrose, S Barrell, R Beale, G Bineva-Todd, D Biswas, R Byrne, S Caidan, P Cherepanov, L Churchward, G Clark, M Crawford, L Cubitt, V Dearing, C Earl, A Edwards, C Ekin, E Fidanis, A Gaiba, S Gamblin, S Gandhi, J Goldman, R Goldstone, PR Grant, M Greco, J Heaney, S Hindmarsh, C F Houlihan, M Howell, M Hubank, D Hughes, R Instrell, D Jackson, M Jamal-Hanjani, M Jiang, M Johnson, L Jones, N Kanu, G Kassiotis, S Kirk, S Kjaer, A Levett, L Levett, M Levi, WT Lu, J I MacRae, J Matthews, L McCoy, C Moore, D Moore, E Nastouli, J Nicod, L Nightingale, J Olsen, N O’Reilly, A Pabari, V Papayannopoulos, N Patel, N Peat, M Pollitt, P Ratcliffe, Sousa C Reis e, A Rosa, R Rosenthal, C Roustan, A Rowan, GY Shin, DM Snell, OR Song, M Spyer, A Strange, C Swanton, JMA Turner, M Turner, A Wack, P A Walker, S Ward, WK Wong, J Wright, M Wu

## Abstract

The emergence of the novel coronavirus SARS-CoV-2 has led to a pandemic infecting more than two million people worldwide in less than four months, posing a major threat to healthcare systems. This is compounded by the shortage of available tests causing numerous healthcare workers to unnecessarily self-isolate. We provide a roadmap instructing how a research institute can be repurposed in the midst of this crisis, in collaboration with partner hospitals and an established diagnostic laboratory, harnessing existing expertise in virus handling, robotics, PCR, and data science to derive a rapid, high throughput diagnostic testing pipeline for detecting SARS-CoV-2 in patients with suspected COVID-19. The pipeline is used to detect SARS-CoV-2 from combined nose-throat swabs and endotracheal secretions/ bronchoalveolar lavage fluid. Notably, it relies on a series of in-house buffers for virus inactivation and the extraction of viral RNA, thereby reducing the dependency on commercial suppliers at times of global shortage. We use a commercial RT-PCR assay, from BGI, and results are reported with a bespoke online web application that integrates with the healthcare digital system. This strategy facilitates the remote reporting of thousands of samples a day with a turnaround time of under 24 hours, universally applicable to laboratories worldwide.

## Introduction

At the end of 2019, an outbreak of pneumonia was observed in Wuhan, China and the pathogen was confirmed as a novel coronavirus strain. The World Health Organization has named the pathogen severe acute respiratory syndrome coronavirus 2 (SARS-CoV-2) [1-3]. The rapidly spreading respiratory tract pathogen led to a pandemic, which together with high hospitalization rates and requirements for critical care, has imposed major challenges to healthcare systems worldwide. Comprehensive and reliable testing is essential to identify the virus in individuals presenting with COVID-19 symptoms in hospital, to guide community interventions that contain the spread, and to perform enhanced surveillance programs of healthcare workers to maintain a workforce to safely deliver care. These requirements have placed an unprecedented demand on the testing capability of all countries. This demand on diagnostic laboratories, coupled with a global shortage of commercial kits and reagents, reduced commercial flights and cargo capacity and international competition for testing resource, has rendered the testing capacity of many countries inadequate to deal with the outbreak effectively.

The Francis Crick Institute (FCI) is a biomedical research institute dedicated to the discovery of biology underlying human health. Situated in central London, an epicenter of the UK pandemic, the FCI elected to repurpose its scientific and technical resource to support the immediate healthcare needs of its partner hospital, University College London Hospital, during the outbreak. Providing an end-to-end pipeline for clinical diagnostic testing of COVID-19, would result in increased testing capacity that could meet local demand, and allow new surveillance programs of healthcare workers, to be implemented. Here we describe the roadmap by which the CRICK COVID-19 Consortium delivered this pipeline, and highlight important considerations to implement similar solutions by research institutes worldwide.

Key to finding a solution was the partnership created between the FCI, a major London healthcare provider together with its clinical virology expertise (University College London Hospitals National Health Services Trust) and an accredited clinical diagnostic laboratory (Health Services Laboratories), forming the CRICK COVID-19 Consortium (CCC). This partnership, effectively removed the barriers of clinical translation, and facilitated rapid implementation of robust end to end testing within ten days under the oversight of an accredited laboratory. Importantly it also allowed resources and expertise to be mobilised to meet local healthcare needs.

A notable strength of the CCC pipeline is that swabs can be either dry or in any proprietary virus transport media (VTM). These are taken at hospital sites or local drive-through stations and submitted to the accredited reporting laboratory before being transferred to the FCI. Specimens are barcode tracked, then proceed immediately to viral inactivation, automated extraction of viral RNA using a series of in-house buffers and RT-PCR to quantify SARS-CoV-2 RNA. Results are accessed through a custom-made online web portal facilitating data to be analysed remotely by a panel of trained reporters, and are returned to the reference laboratory. The speed and precision of the pipeline permits the reporting of thousands of samples/day, adopts processes that are widely used by many research laboratories worldwide, and is free from dependence on supply chain constraints.

## Methods

### Formation of a consortium

To facilitate the accelerated establishment of the CRICK COVID-19 Consortium pipeline within a fortnight, we partnered with an accredited diagnostic provider (Health Services Laboratories, HSL) and a healthcare trust and its Clinical Virology department (University College London Hospital, UCLH). HSL evaluated and validated our RT-PCR assay against their diagnostic SARS-CoV-2 N gene assay and acted as the reference laboratory during the conception of the CCC pipeline. UCLH provided access to barcoded swabs pre-booked onto our Laboratory Information Management System (LIMS) to enable tracking from sample receipt through to result reporting. In urgent response to the clinical need, formation of these partnerships was vital to drive the speed of pipeline setup in our central London research laboratory.

### Accreditation and Governance

We have worked in partnership with HSL - the UCLH UKAS accredited lab, who already had a COVID-19 test in scope. All samples are received and communicated by HSL, under their accreditation and the CCC assay was validated against the existing HSL test. Given the urgent timeframe required to implement testing, it was not possible to secure clinical laboratory accreditation for the FCI, to an appropriate standard (ISO 15189:2012; US equiv. CAP/CLIA). However, full measures were taken to ensure that the CCC test was evaluated, verified and performed for diagnostic use in an environment that adhered to equivalent international standards, overseen and audited by HSL. These measures, were implemented under the advice and oversight of registered professionals from existing nearby ISO accredited Medical Laboratories, and included writing and following clinical diagnostic Standard Operating Procedures (SOPs) for every stage of the pipeline from sample reception, processing to result reporting by qualified clinical scientists prior to results being sent to patients by HSL. Additional SOPs were followed for sample storage, disposal of materials, batch certification of reagents and incident reporting. Appropriate risk assessments, training and competency assessment procedures were established and documented. Record sheets were created to document the receipt, batch acceptance testing, and start/end of use dates for key reagents and consumables. An inventory of all key equipment was compiled which, where appropriate, included details of service and calibration records. Systems were also established for the control of all key documents (version implementation, distribution and acknowledgement), audit trailing (what samples were tested when, by whom, with what equipment and using which consumable/reagent batches), and the recording of all untoward incidents/issues (thus facilitating appropriate investigation, rectification and recurrence prevention). Samples were barcoded and tracked using the Crick Clarity LIMS system. All key documents are available at https://www.crick.ac.uk/research/covid-19/covid19-consortium. NHS Governance was extended by a specific memorandum of understanding for diagnostic PCR testing during the pandemic between UCLH and the Crick Institute and was enabled by NHS England. Assurance of the pipeline was performed in collaboration with GenQA, following their checklist for non-accredited laboratories, and the lab and CCC workflow were inspected by a qualified UKAS assessor against the GenQA guidelines to verify compliance to, IS 015189 equivalent standard.

### CCC test procedure

The protocol described below is for 94 samples processed through the CCC pipeline, with a positive and negative control added prior to RT-PCR. The procedure can be scaled up as required to increase throughput. The CCC pipeline is illustrated in Figure 1A. The specific reagents and requirements for each step of the entire pipeline are detailed in Supplementary Methods 1-16.

### Sample receipt

All specimens are swabs submitted in a collection tube either dry or in presence of Virus Transfer Medium (VTM). All samples are barcoded prior to arrival at the test laboratory and initially unpacked and tracked using the specimen management tool Tube Tracker (Cerebros Medical Systems). Following visual inspection of sample integrity, samples are scanned onto our internal LIMS. Incorrect or leaky samples are returned or disposed of in accordance with our internal safety regulations and risk assessments (See Supplementary Method 1). Using the CCC pipeline, 94 samples can be processed by 4 operators within 25 minutes.

### Virus inactivation

This procedure is carried out within a Class I or II safety cabinet by trained authorised personnel only, with personal protective equipment (PPE) at all times. After further inspection of sample integrity, the swab is removed and, if the sample contains VTM, 100μL of each specimen is transferred to a barcoded 2-ml screw cap tube prefilled with 1ml of 5M guanidinium thiocyanate L6 virus inactivation buffer ((Boom, Sol et al. 1990)) (See Supplementary Methods 11, 12, 13, 14 and 15). If the swab is submitted dry, it is immersed in inactivation buffer [4]. Following transfer, barcodes of both the collection tube and the inactivated viral sample are visually checked to confirm identity. Only one sample is processed at a time to avoid any risk of sample swapping. Liquid waste is decontaminated with 10% Surfanios. Inactivated individual viral samples are then moved to a separate area for RNA extraction. 94 specimens can be inactivated by 4 operators in 47 minutes using the method detailed in Supplementary Method 2.

### RNA Extraction

The first step consists of transferring 150μL of inactivated lysate from each individual sample tube to a Nunc 96-DeepWell polypropylene storage plate to allow batch processing. This is performed by one operator on a Hamilton STAR or STARlet liquid handling platform where all individual barcodes are automatically scanned ensuring errorless sample tracking (Supplementary Method 3). The RNA extraction procedure, adapted from Rohland et al. [5], uses pre-prepared batch-certified binding buffer (BB) and silica-coated magnetic beads (G-Biosciences) to extract RNA using a home-made guanidine hydrochloride solution. The RNA extraction process is automated on a Biomek FX liquid handling platform (Beckman Coulter). This requires one operator to set up, takes approximately 49 minutes and results in RNA eluted in a 96-well barcoded plate (Supplementary Methods 4, 9, 10).

### RT-PCR

10μL of extracted RNA are used for the RT-PCR assay. Primers and fluorescent probes in the BGI kit are designed against the highly conserved O, RF 1ab region of SARS-CoV-2. The primer sequences are located within the region of SARS-CoV-2 O, RF 1ab 3000-4000; (there is only one unique site of the SARS-CoV-2 genome for qPCR in the BGI kit). The oligonucleotide probes have a FAM fluorophore reporter attached to the 5’ end. A specific primer and probe set for human beta-actin is included as an internal reference with the VIC fluorophore as a reporter at the 5’ end. The use of the internal control permits swab failures to be differentiated from SARS-CoV-2-negative specimens which is critical when dealing with dry and wet swabs. Each batch of 94 samples is run with a positive control provided in the kit and a sample-free eluate as blank control. Each batch of samples is run in duplicate. The RT-PCR protocol using the ABI QuantStudio 3 takes 88 min to complete and allows the quantitative assessment of SARS-CoV-2 in specimens. A reference plate is run on alternate days as a quality control step. (Supplementary Methods 5, 6, and 7).

### N gene assay

The CCC test was validated against the N gene assay developed by the reference laboratory (Health Services Laboratories, HSL) [6]. The N gene assay is a high throughput RT-PCR assay that runs on the Hologic Panther Fusion platform using the Open Access function which allows the use of custom primers and probe targeting the nucleocapsid (N) gene of SARS-CoV-2 and includes Hologic internal control primers and probe; N gene Taq1 (TCTGGTAAAGGCCAACAACAA), N gene Taq2 (TGTATGCTTTAGTGGCAGTACG) and N gene probe with 5’ Fam (CTGTCACTAAGAAATCTGCTGCTGAGGC). The N gene assay was previously validated against a SARS-CoV-2 assay from Public Health England run at the Colindale reference laboratory.

### Assessment of data quality

An initial quality control assessment of the integrity of the run, is performed to establish whether both internal and external controls have passed the set criteria; for the blank control, the Ct values at FAM (COVID-19) and VIC (internal control) channels must be in the negative ranges (>37.0 or no data available for FAM and >35.0 or no data available for VIC). These criteria are in line with the manufacturers instructions (https://www.bgi.com/us/wp-content/uploads/sites/2/2020/03/EUA-Real-Time-Fluorescent-SARS-2019-nCoV-BGI-IFU.pdf); for the positive control, the standard curves at channel FAM and VIC channels must be S-shape with Ct values not higher than 37.0 and 35.0 respectively. These requirements should be met on the individual plate otherwise the entire plate is deemed invalid. The assessor must have experience in clinical reporting of RT-PCR data and be registered with the Health and Care Professionals Council (HCPC) as Biomedical Scientist or Clinical Scientist, or be a fellow of the Royal Colege of Pathologists. The assessor should also ensure there is a complete audit trail for all sample processing steps. The data are then exported for remote clinical interpretation. For a test sample, a COVID-19 Ct <37.0 with typical exponential amplification regardless of any internal control (IC) value gives an automated POSITIVE result. A signal of COVID-19 ‘undetermined’ or Ct >37.0 with IC<35.0 gives an automated negative result. A COVID-19 ‘undetermined’, or Ct>37.0 with IC>35.0 gives an automated ‘failed sample’ result as this may represent an inadequate swab. The thresholds set for the CCC test are based on the manufacturer’s guidelines (BGI) and are consistent with our control data. All results are uploaded to the pathology LIMS system as a .xls file export, with logic created to generate automated results in line with the rules stated above.

### Online web reporting

Following the initial quality control performed by the first reporter, a second reporter is able to access the data for each duplicated run through a web application (github access available on request; Supplementary Method 8), evaluate and upload the results as an excel file. The roles of the second reporter include checking of positive and negative controls as described above and confirming the acceptance criteria have been passed, the overall plate curves suggest appropriate PCR amplification and ensuring there is a complete audit trail for all sample processing steps. The second reporter manually sets the threshold above the level of background non-exponential amplification and exports data to the reference laboratory. Positive, negative or sample failure results will be generated automatically based on the Ct values or comments in the .xls results that are uploaded to the reporting system in the reference laboratory. Logic in the online reporting system will ensure repeat samples are requested for any discordant results across the duplicate runs. This step of the pipeline allows results from 94 patients to be generated within 20 minutes. Clinical reporting and authorisation rules are implemented: a) clinical comments are automated with each report and b) mobile phone texts are subsequently used to alert the individual that was tested. For Healthcare Workers (HCWs) this is particularly useful as illustrated below with the texts used at reporting: If DETECTED: Text: “Your COVID-19 test is POSITIVE; self-isolate for at least 7 days.” If NOT Detected: Text: “Your COVID-19 test is NEGATIVE; consider return to work following local Occupational Health Policy” If INVALID:Text: “Your COVID-19 test is INVALID; we recommend a REPEAT swab”. (Figure 1B and Supplementary Method 8).

**Figure 1).**
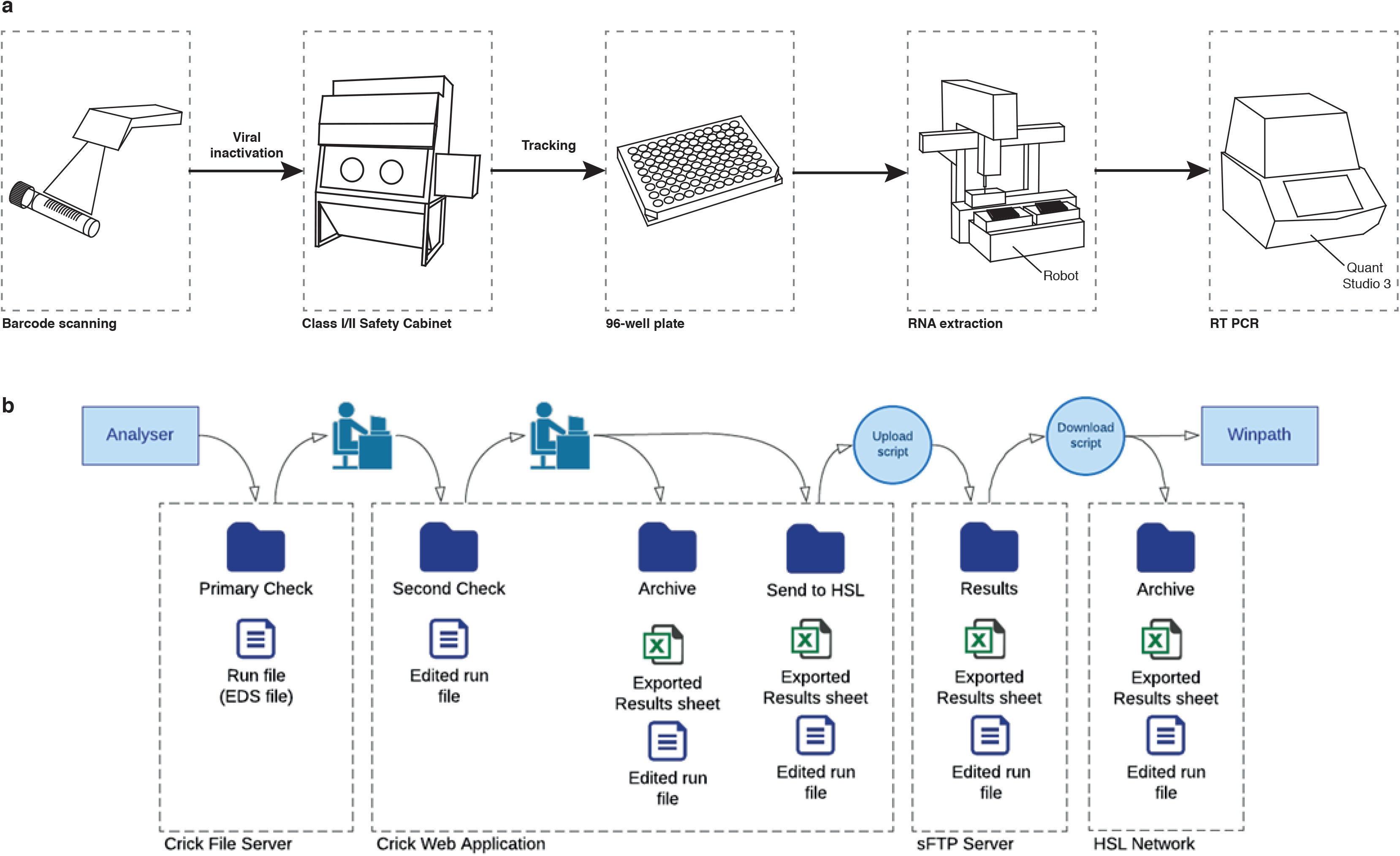
Schematic of the CRICK COVID-19 Consortium test and reporting pipelines A) For the CRICK COVID-19 Consortium test, specimen barcodes are scanned at sample reception, prior to viral inactivation in a Class I or II safety cabinet, processing through RNA extraction using an in-house protocol and RT-PCR testing using a commercial kit (BGI). **B)** CRICK COVID-19 Consortium reporting pipeline. Test results are reported through a custom-made remote web application, allowing remote clinical scientists and pathologists working outside of the institute to authorise reports, in line with the established SOP.

### Barcode tracking

Our streamlined pipeline ensures every sample is individually tracked from hospital swabs, to 2-ml viral inactivation samples, to aliquoting into barcoded 96-well plates, to 96-well plates for RNA extraction, a further 96-well barcoded plate for RT-PCR down to final remote reporting. Sample requests are sent from our reference laboratory’s WinPath system and are uploaded to a secure FTP site to create an entry in the GenoLogics ClarityLIMS sample tracking database system and the swab specimens are registered into a sample queue. Samples received into our sample reception area are scanned to update the database and critically assessed. If they are damaged, the samples are scanned out so as to mark them as removed from further processing. Samples are subjected to viral inactivation and transferred to barcoded plates for the subsequent steps of RNA extraction and RT-PCR. ClarityLIMS is commercially licensed web-based sample tracking software installed on-premises. The application is integrated into the Francis Crick internal system. Other sample management systems can be used but configuration and integration into existing pipelines will be required.

## Results

### CCC test validity

The CCC test was bench-marked against the reference laboratory N gene assay using 56 current clinical samples processed in parallel using the Hologic Panther platform. 27 specimens were processed in duplicate through the CCC pipeline from 150μL inactivated virus, through RNA extraction and RT-PCR (Figure 2A) and a second batch of 29 clinical specimens was processed independently (Figure 2B), all with a parallel test of the same sample processed through the N gene assay on the Hologic Panther platform. As the reference laboratory assay does not contain a human internal control, there was no mechanism for testing ‘swab failures’ with the N gene assay. As part of the validation, 3 untested ‘blank’ swabs were submitted, yielding a negative result by the reference laboratory assay, but due to the absence of signal in the human internal control were classified as swab fails by the CCC test (Figure 2C). 5 of the clinical samples submitted were classified as ‘low level positives’ by the N gene assay with Ct values of >38.0. These 5 samples were all repeated using the N gene assay and again returned high Ct values and are therefore classified as borderline positives (Supplementary Figure 1). Overall, the diagnostic sensitivity of the CCC test is 92.86% with a specificity of 100% and a high degree of accuracy in the detection of SARS-CoV-2.

**Figure 2).**
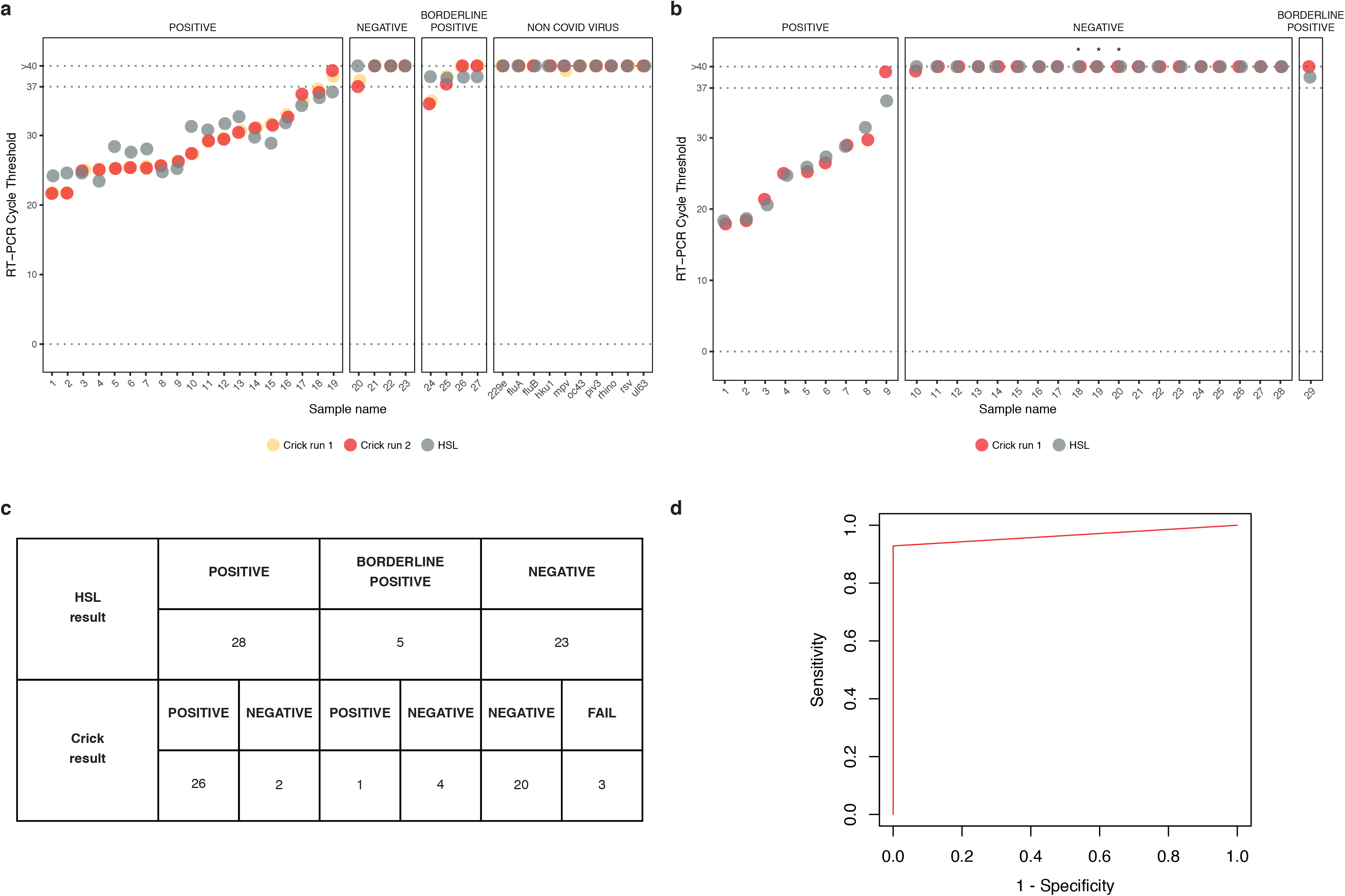
CRICK-COVID-19 Consortium RT-PCR test validity and specificity. A) Dot plot demonstrating the COVID-19 Ct values obtained from 2 repeats of processing 37 samples through the CCC test (red and orange dots) alongside Ct values derived from the reference laboratory using the N gene assay (gray dots). The clinical call from the reference laboratory is indicated above the boxes; POSITIVE, NEGATIVE, BORDERLINE POSITIVE or NON COVID VIRUS. All repeats were independently re-extracted from the original inactivated virus solution. Ct values of ‘undetermined’ are plotted as “>40” for illustrative purposes. This set of 37 includes 27 current clinical samples taken for diagnosis of COVID-19 and a further 10 samples classified as Non-COVID virus from patients known to be infected with other viruses such as Flu A, Flu B, RSV, rhinovirus, metapneumovirus, parainfluenza virus. **B)** Dot plot demonstrating the COVID-19 Ct values obtained from a repeat (1 test performed at the research institute using the BGI kit, 1 performed at the reference laboratory using the N gene assay) of processing an independent batch of 29 samples through the RT-PCR assay. All repeats were independently re-extracted from the original inactivated virus solution. Ct values of ‘undetermined’ are plotted as “>40” for illustrative purposes. CCC ‘failed’ swabs, with negative internal control signal, are marked with the ‘*’ symbol. **C)** Table to summarise the concordance of the CCC test with the reference laboratory results for the 56 COVID-19 testing specimens from A and B. **C)** ROC curve illustrating CCC test specificity and sensitivity. The positive predictive value is 100% and the negative predictive value is 90.9%.

### CCC test specificity

Cross-reactivity of the SARS-CoV-2 RT-PCR probes in the BGI kit was tested by performing the CCC RT-PCR test using specimens from 10 patients known to be infected with other viruses such as flu, RSV, rhinovirus, metapneumovirus and parainfluenza virus (Figure 2A). Specimens from patients with other viruses resulted in COVID-19 Ct values ‘undetermined’ or above 37.0 in the assay (negative result) demonstrating a high degree of assay specificity against SARS-CoV-2.

The specificity of the SARS-CoV-2 RT-PCR assay was further tested against RNA extracted from human cell lines (1:1 mixture of A549 and, HT 1080 cell lines) compared to a single positive control sample. Only the positive control well displayed SARS-CoV-2 amplification in the RT-PCR. All human cell lines were negative at 1:125 dilution (Supplementary Figure 2A). We next monitored our test specificity during live pipeline runs by assessing the Ct values from the SARS-CoV-2 amplification for wells containing elution buffer and RT-PCR master mix only (non-sample wells). Twenty-four out of twenty-five non-sample wells processed in our initial 3 live runs had an undetermined Ct value reflecting the absence of an amplified product in these wells (Supplementary Figure 2B). 48 samples of water were tested alongside a positive control to assess the occurrence of inherent late-cycle amplification or non-specific signal from using this RT-PCR kit. Late Ct signals for the internal control (>35) and COVID-19 (>37) targets were observed in 2 of 48 and 1 of 48 wells respectively (Supplementary Figures 3A and B). When the experiment was repeated using guanidinium inactivation buffer and RNA elution buffer, the signal was present in 8 out of 48 and 2 out of 48 wells respectively (Supplementary Figures 3C and D). No overlap was seen for the 2 targets, confirming that late signals did not represent contamination from positive control. The Ct values reported in the guanidinium and RNA elution buffer controls support the thresholds set for calling positive, negative and failed samples as documented in the manufacturer’s instructions. The reproducibility of the kits used for the CCC test was determined by comparing two different batches of RT-PCR kit reagents (BGI). Four serial dilutions of the positive control from the kit were assessed in columns of a 96 well plate alongside 2 columns of negative controls. Both batches gave linear results following serial dilution. For the negative control wells, both batches gave similar degrees of late stage amplification; AUTO analysis yielded Ct’s above 38 for COVID-19 and above 37 for the internal control (Supplementary Figure 4).

**Figure 3).**
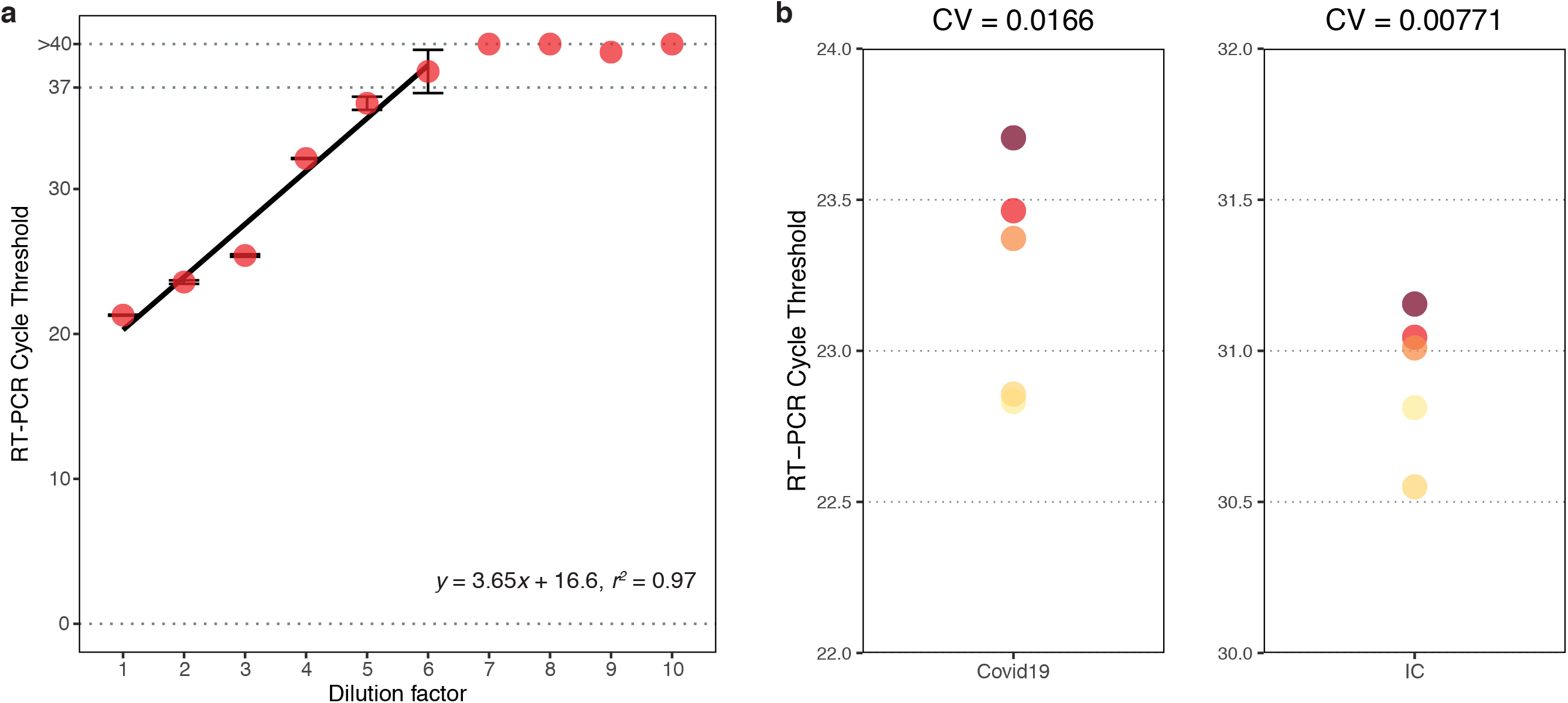
CRICK COVID-19 Consortium test sensitivity and reproducibility A) SARS-CoV-2 titration curve. Ten-fold serial dilutions of patient samples were carried out by the reference laboratory. Following RNA extraction, RT-PCR was performed to determine the linearity of the detection process using the BGI kit. **B)** Dot plot showing assay precision. Five aliquots of one COVID-19 positive sample were processed through independent repeats of RNA extraction and RT-PCR using the BGI kit to determine the precision of the CCC test. Data are shown for COVID-19 target (left) and internal control target (right). CV = coefficient of variation.

### CCC test sensitivity

In an effort to ascertain the CCC test sensitivity, particularly relevant to specimens with low viral concentration, a linearity analysis was performed by testing 10-fold serial dilutions of RNA extracted from a COVID-19 positive patient followed by RT-PCR. Starting with a Ct value of 21.3 in the original sample, a linear response was observed to 10^6^ dilution from the original RNA eluate (Figure 3A).

### CCC test reproducibility

The reproducibility of the CCC test was measured using a sample from a patient with confirmed COVID-19 diagnosis. The sample was diluted 100 fold in viral inactivation buffer and individual aliquots were subject to 5 independent repeats of the CCC pipeline through RNA extraction and RT-PCR to determine Ct values (Figure 3B). The coefficient of variation for CCC test precision was calculated as 0.0166.

## Discussion

Here we describe how a partnership between a biomedical research institute, clinical virologists and a diagnostic laboratory can be formulated to meet a healthcare need during a viral pandemic. We present a sensitive, specific and high-throughput pipeline (24hr turnaround) for detection of SARS-CoV-2 RNA in individuals with suspected infection. A number of approaches have been established to detect SARS-CoV-2 in specimens and several tests are currently available that use different approaches to inactivate the virus as well as RNA extraction-free approaches [6-9]. We discuss the key considerations that informed the specific approaches adopted in the CCC pipeline.

Viral inactivation is performed in a containment level 3 suite, compatible with our local, CL 3 capacity and trained staff at FCI. With appropriate risk assessments unique to different research institutions or with swabs being inactivated prior to transportation, the pipeline could be performed in containment level 2 facilities, adapting to more laboratories worldwide (Supplementary Method 16). Protocols also exist for alternative viral inactivation methods using heat further demonstrating potential for CCC pipeline applicability where availability of guanidinium may be limited.

The CCC test uses a series of home-made buffers for automated RNA extraction and routine RT-PCR using a kit from BGI which circumvents dependence on reagents that may be in short supply during a pandemic. At the time of writing the most important bottleneck in performing PCR tests for COVID-19 detection is the shortage of kits for RNA extraction. We developed an in house RNA extraction protocol using magnetic silica beads from G-Biosciences, and we have also validated our assay with SeraSil Mag 400 beads (GE Healthcare/Cytiva), which can serve as a reliable substitute. RNA extraction using silica is based on the protocol developed by Boom et al. over 30 years ago [10]. In the Boom method, concentrated guanidinium thiocyanate serves as virus and RNase inactivation agent and promotes binding of nucleic acids to silica. An alternative method, solid-phase reversible immobilization (SPRI), takes advantage of nucleic acid binding to magnetic beads coated with carboxyl groups in the presence of polyethylene glycol and salt [11]. We have tested Beckman RNAclean XP SPRI magnetic beads (Beckman) and found them compatible with our virus inactivation solution L6; viral RNA could be purified following manufacturer’s recommendations. Moreover, protocols exist for the production of either type of magnetic beads from inexpensive and accessible starting materials [12]. Therefore, we designed a pipeline that uses common reagents and is automatable on widely available liquid handling platforms allowing its implementation in a large number of biomedical laboratories with suitable robotic platforms that can be reprogammed for this use. The universal applicability of this approach could allow a resilient response to future critical events even in countries where particular resources may be limited.

Selection of an appropriate PCR assay for detection of SARS-CoV-2, the BGI kit, was based on (i) our accredited laboratory with a ready set of validation data and experience with the FDA U.S. Food and Drug Administration approved assay and (ii) a guaranteed supply chain for the assay kit in the face of falling demand in China, and growing demand in the US (for US suppliers). The verification steps of the CCC pipeline allowed us to compare the BGI kit with the in-house developed N gene assay also reported here. SARS-CoV-2 is an enveloped, positive sense, single stranded RNA virus. In common with other coronaviruses, non-structural proteins including the RNA dependent RNA polymerase are encoded within O, RF 1a/b at the 5’ end of the genome. Structural proteins spike (S), envelope (E), membrane (M) and nucleoprotein (N) are produced from a set of nested subgenomic mRNAs co-terminal with the 3’ end of the genome. N is situated at the 3’ terminus and is encoded by all subgenomic and genomic RNAs (reviewed in Sawicki et al 2007) [13]. The primers used in the CCC test target O, RF 1a, enabling detection of full length genomic and antigenomic RNA, whereas the N gene assay also targets the abundant subgenomic RNAs. Consistent with this, the N gene assay is slightly more sensitive than the CCC assay at the limits of detection (Figure 2A and 2B). Sample timing and adequacy are likely to be more important determinants of false negatives than qPCR sensitivity [14]. The CCC test includes a control for cellular RNA (beta-actin), which serves as a partial proxy for sample adequacy. Although high sensitivity at the assays limits of detection could impact identification of low levels of viral shedding beyond the assay’s limits of detection, these samples are unlikely to be producing infectious virus [15]. Moreover, PCR testing in mildly symptomatic HCWs is not recommended beyond 5 days from onset and therefore we believe the BGI PCR exhibits adequate sensitivity for current clinical algorithms. The high throughput RT-PCR assay in 96-well plate format has the potential to screen thousands of samples per day and can be scaled up to 384-well format with further optimisation.

Medical laboratory accreditation is held to the standard of The International Standards Organisation (ISO) 15189:2012 across the world, with the exception of the USA which operates to the standard of Clinical Laboratory Improvements Amendment (CLIA) certification and College of American Pathologists (CAP) accreditation. Laboratories are assessed for compliance to ISO or CLIA/CAP standard by a national awarding body; in the United Kingdom this body is the UK Accreditation Service (UKAS). The CCC was established in partnership with Health Services Laboratories (HSL), an existing UKAS-accredited clinical laboratory, to deliver COVID-19 testing. The process for acquiring accreditation, and the typical assessment time span and rules for extending existing ISO or CLIA/CAP scope to partnering institutions will vary between countries and wherever possible clinical accreditation should be sought by research institutions seeking to establish clinical testing. Whilst pursuing this process, our approach has been to implement processes in line with international accreditation standards, and those processes remain under the supervision of our partner accredited laboratory.

Health information systems such as the EPIC electronic medical record used at UCLH interface with laboratory information management systems such as WinPath to enable sample barcodes to be associated with patient hospital numbers. The pipeline set up uses a custom-made reporting web application compatible with remote reporting. This allows multiple trained reporters to access anonymised data through a portal from home, particularly advantageous in a pandemic. This facilitates an accelerated turn-around for results of >3000 samples within 24 hours.

The potential advantanges of implementing a clinical diagnostic pipeline in research laboratories are clear: a significant increase in capacity for testing, and the ability to adopt flexilbe and agile approaches to testing in the face of global constraints. Our experience in implementing mass scale testing within the CRICK COVID-19 Consortium has taught us invaluable lessons for the wider academic community first, diagnostic testing to clinical standards can be successfully achieved through partnership and guidance from a clinical diagnostic laboratory, second, the choice of techniques and approaches should be adapted to the local resource, and staff expertise, already existing within a research laboratory; and third, the scale and implementation of testing should be aligned with the healthcare needs and demands of the local population.

**Supplementary Figure 1).**
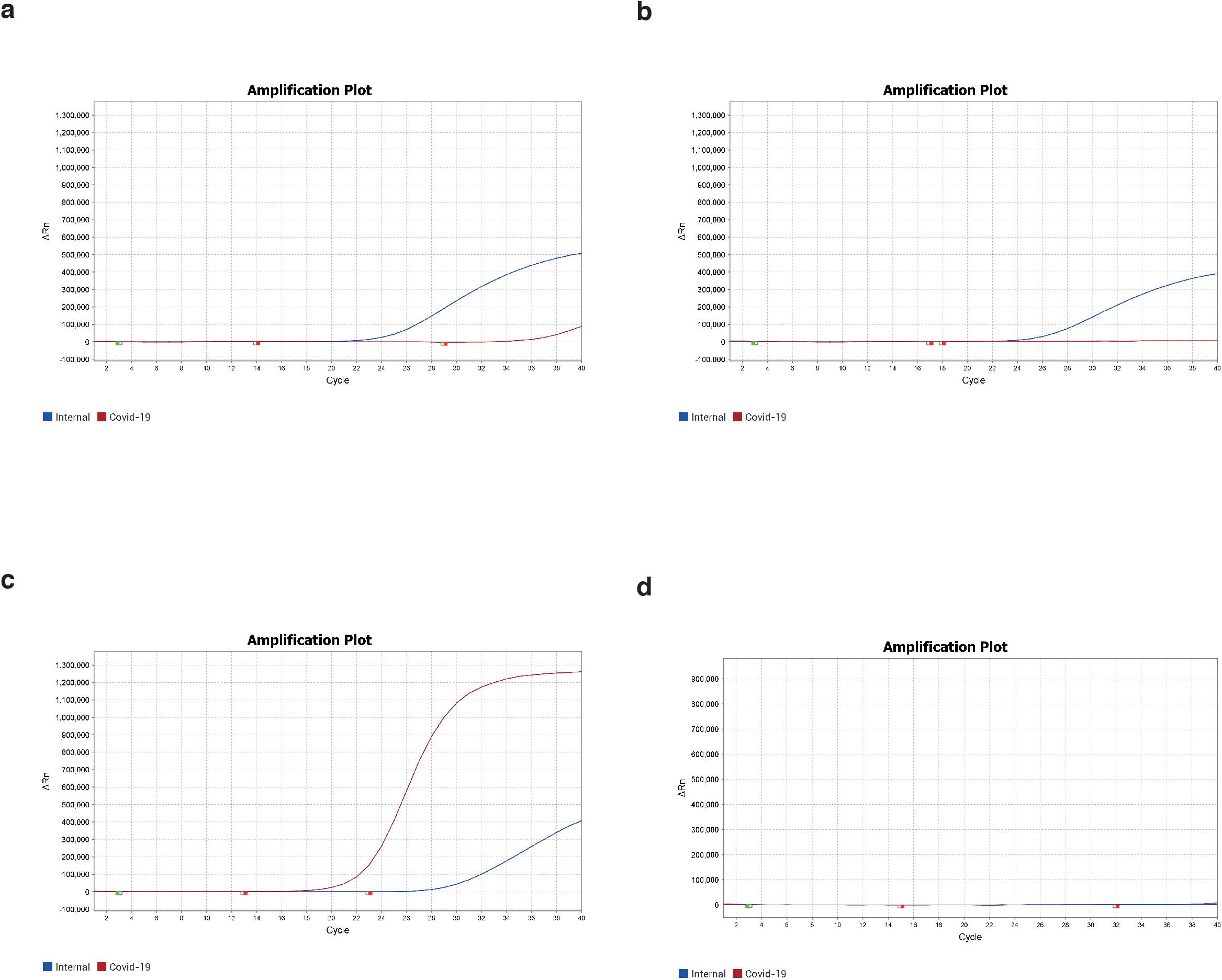
Example amplification plots from the CRICK COVID-19 Consortium test. Illustrations of results that were called **A)** Positive by the reference laboratory, negative by the CCC; **B)** Borderline positive by the reference laboratory, negative by the CCC; **C)** Positive by both the reference laboratory and the CCC; **D)** Negative by the reference laboratory, failed by the CCC.

**Supplementary Figure 2).**
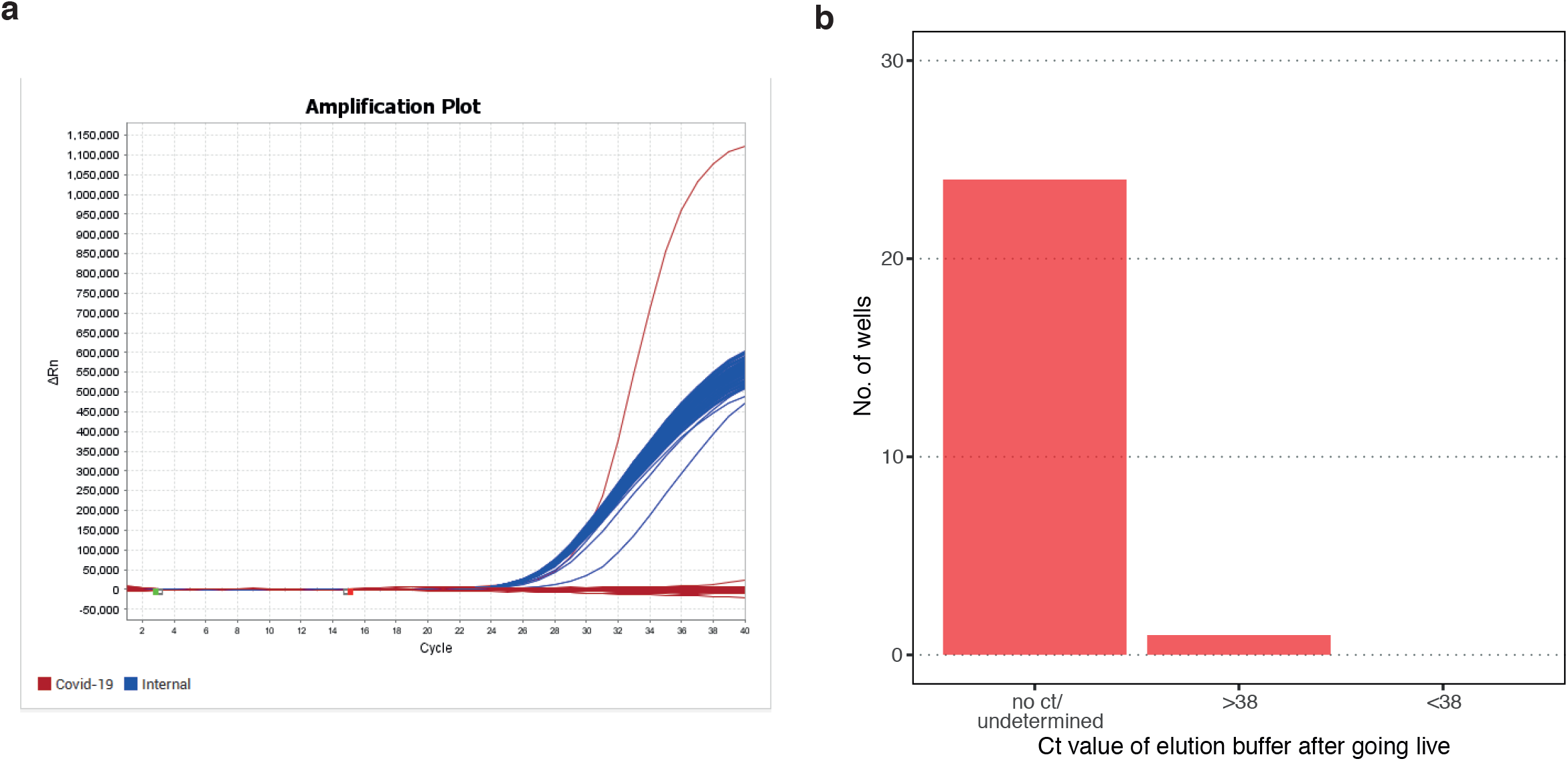
CRICK COVID-19 Consortium test specificity A) CRICK COVID-19 assay specificity using RNA extracted from cell lines. RNA extracted from human cell lines (95 wells) were analysed alongside 1 well of positive control RNA. Only the positive control well displayed a COVID-19 amplification signal (red). The average internal control Ct for human cell line RNA (blue) is 25.55±0.15. **B)** CRICK COVID-19 Consortium assay specificity in “non-sample” wells during “live” runs. For specificity assessment, COVID-19 Ct values from ‘non-sample’ wells (elution buffer + PCR master mix only) were determined during live runs. 24/25 wells showed no detectable Ct value (n=3).

**Supplementary Figure 3).**
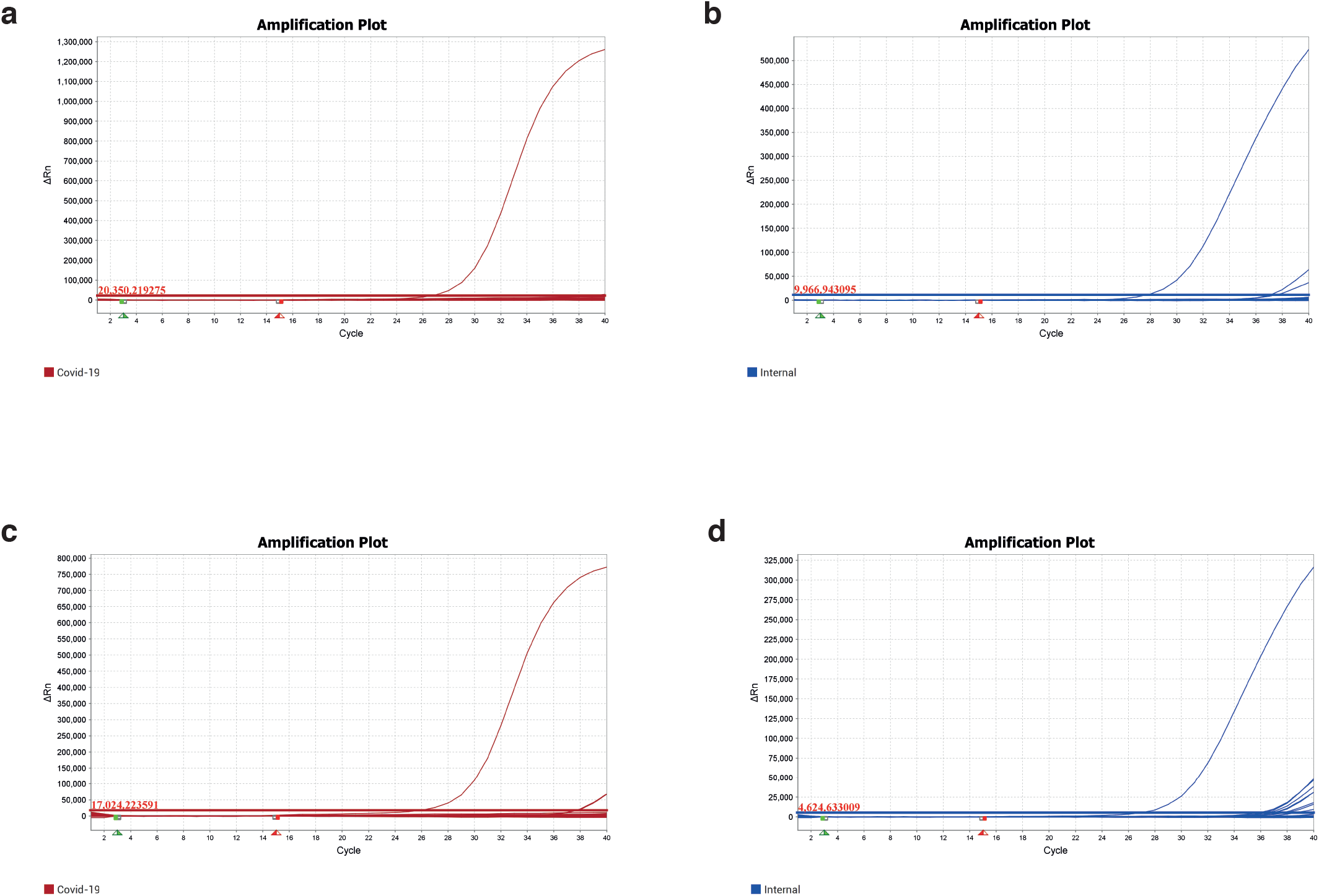
CCC test specificity determined using water or guanidinium buffer and RNA elution buffer. Amplification plots derived from a plate of 48 no template controls (water), plus one positive control well. Late-rising curves above the manually set threshold occur at Ct>39.0 in one well for COVID-19 **(A)** and 2 wells at Ct>36.0 for internal control **(B)**. There is no overlap between the well giving late non-specific COVID-19 signal and the 2 wells showing late internal control signal. Amplification plots derived from a plate of 48 no template controls (guanidinium + RNA elution buffer), plus one positive control well. Late-rising curves above the manually set threshold occur at Ct>37.0 in 2 wells for COVID-19 **(C)** and 8 wells out of 48 at Ct>35.0 for internal control **(D)**. There is no overlap between the 2 wells giving late non-specific COVID-19 signals and the 8 wells showing late internal control signals. Cts of late rising curves reduce by 1 -2 cycles from water control to guanidinium + RNA elution buffer.

**Supplementary Figure 4).**
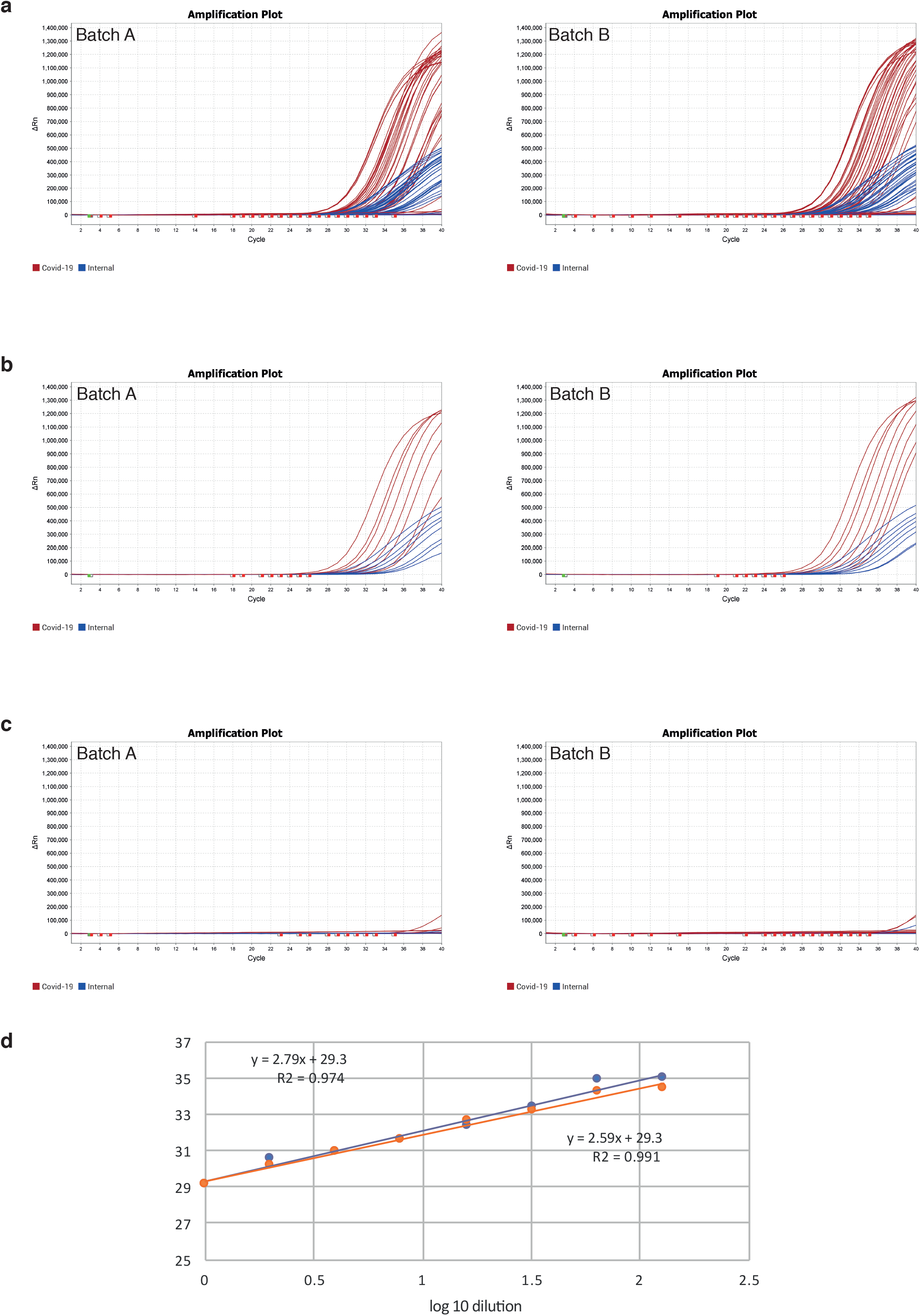
CCC test BGI kit batch reproducibility assessment. A) Amplification plots from a 96 well plate with four columns of 10 fold serial dilutions and 2 columns of negative control run in parallel with 2 different batches of RT-PCR kit from BGI. **B)** Amplification plots illustrated for one column from each kit, which is a 2-fold dilution of positive control down the column. **C)** Amplification plots of 2 columns of negative control loaded wells demonstrating similar numbers of late stage amplification. **D)** Titration curve calculated from data in A for the two batches (blue, batch A, orange batch B).

## Data Availability

Extended protocols are included in the Supplementary Methods

## Acknowledgements

This work was supported by the Francis Crick Institute that receives its core funding from Cancer Research UK (, FC 001169,, FC 001078), the UK Medical Research Council (, FC 001169,, FC 001078), and the Wellcome Trust (, FC 001169,, FC 001078). C.S. is Royal Society Napier Research Professor and is also funded by the Breast Cancer Research Foundation (BCRF). E.N. receives research funding from the NIHR, MRC, GSK and H2020. S. Gandhi is an MRC Senior Clinical Fellow.The authors wish to thank Heather Ringrose for support with the Hamilton liquid handling workstation.

## References

1. Jiang, S., L. Du, and Z. Shi, An emerging coronavirus causing pneumonia outbreak in Wuhan, China: calling for developing therapeutic and prophylactic strategies. Emerg Microbes Infect, 2020. 9(1): p. 275–277.

2. Zhou, P., et al., A pneumonia outbreak associated with a new coronavirus of probable bat origin. Nature, 2020. 579(7798): p. 270–273.

3. Zhu, N., et al., A Novel Coronavirus from Patients with Pneumonia in China, 2019. N Engl J Med, 2020. 382(8): p. 727–733.

4. Moore, C., et al., Dry cotton or flocked respiratory swabs as a simple collection technique for the molecular detection of respiratory viruses using real-time NASBA. J Virol Methods, 2008. 153(2): p. 84–9.

5. Rohland, N., et al., Extraction of highly degraded DNA from ancient bones, teeth and sediments for high-throughput sequencing. Nat Protoc, 2018. 13(11): p. 2447–2461.

6. Grant, P.R., et al., Extraction-free COVID-19 (SARS-CoV-2) diagnosis by RT-PCR to increase capacity for national testing programmes during a pandemic. bioRxiv, 2020: p. 2020.04.06.028316.

7. Fomsgaard, A.S. and M.W. Rosenstierne, An alternative workflow for molecular detection of SARS-CoV-2 - escape from the NA extraction kit-shortage. *medRxiv*, 2020: p. 2020.03.27.20044495.

8. Pfefferle, S., et al., Evaluation of a quantitative RT-PCR assay for the detection of the emerging coronavirus SARS-CoV-2 using a high throughput system. Euro Surveill, 2020. 25(9).

9. Yu, F., et al., Quantitative Detection and Viral Load Analysis of SARS-CoV-2 in Infected Patients. Clin Infect Dis, 2020.

10. Boom, R., et al., Rapid and simple method for purification of nucleic acids. J Clin Microbiol, 1990. 28(3): p. 495–503.

11. DeAngelis, M.M., D.G. Wang, and T.L. Hawkins, Solid-phase reversible immobilization for the isolation of PCR products. Nucleic Acids Res, 1995. 23(22): p. 4742–3.

12. Oberacker, P., et al., Bio-On-Magnetic-Beads (BOMB): Open platform for high-throughput nucleic acid extraction and manipulation. PLoS Biol, 2019. 17(1): p. e3000107.

13. Sawicki, S.G., D.L. Sawicki, and S.G. Siddell, A contemporary view of coronavirus transcription. J Virol, 2007. 81(1): p. 20–9.

14. Wikramaratna, P., et al., Estimating false-negative detection rate of SARS-CoV-2 by RT-PCR. medRxiv, 2020: p. 2020.04.05.20053355.

15. Wolfel, R., et al., Virological assessment of hospitalized patients with COVID-2019. Nature, 2020.

